# Attitudes towards vaccines and intention to vaccinate against COVID-19: Implications for public health communications in Australia

**DOI:** 10.1101/2021.09.12.21263158

**Authors:** Joanne Enticott, Jaskirath Gill, Simon Bacon, Kim Lavoie, Dan Epstein, Shrinkhala Dawadi, Helena Teede, Jacqueline Boyle

## Abstract

**Objective:** To examine SARS-CoV-2 vaccine confidence, attitudes and intentions in Australian adults.

**Methods:** Nationwide survey in February-March 2021 of adults representative across sex, age and location. Vaccine uptake and a range of putative drivers of uptake, including vaccine confidence, socioeconomic status, and sources of trust, were examined using logistic and Bayesian regressions for vaccines generally and for SARS-CoV-2 vaccines.

**Results:** Overall 1,166 surveys were collected from participants aged 18-90 years (mean 52, SD of 19). Seventy-eight percent reported being likely to receive a vaccine against COVID-19. Higher SARS-CoV-2 vaccine intentions were associated with: increasing age (OR: 1.04 95%CI [1.03-1.044]), being male (OR: 1.37, 95% CI [1.08 – 1.72]), residing in the least disadvantaged area quintile (OR: 2.27 95%CI [1.53 – 3.37]) and a self-perceived high risk of getting COVID-19 (OR: 1.52 95% CI [1.08 – 2.14]). However, 72% of participants did not believe that they were at a high risk of getting COVID-19. Findings regarding vaccines in general were similar except there were no sex differences. For both the SARS-CoV-2 vaccine and vaccines in general, there were no differences in intentions to vaccinate as a function of education level, perceived income level, and rurality. Knowing that the vaccine is safe and effective, and that getting vaccinated will protect others, trusting the company that made it and getting vaccination recommended by a doctor were reported to influence a large proportion of the study cohort to uptake the SARS-CoV-2 vaccine. Seventy-eight percent reported the intent to continue engaging in virus-protecting behaviours (mask wearing, social distancing etc.) post-vaccine.

**Conclusions:** Seventy-eight percent of Australians are likely to receive a SARS-CoV-2 vaccine. Key influencing factors identified in this study (e.g. knowing that the vaccine is safe and effective, getting a doctor’s recommendation to get vaccinated) can be used to inform public health messaging to enhance vaccination rates.

**Strengths and limitations of this study:**

- This research captured a large, representative sample of the adult Australian population across age, sex, location, and socioeconomic status.
- We have self-reported Australian uptake intentions and attitudes on general vaccines and COVID-19 vaccine, and intent to continue engaging in virus-protecting behaviours (mask wearing, social distancing etc.) post SARS-CoV-2 vaccine.
- We examine a range of drivers and factors that may influence intent to get the SARS-CoV-2 vaccine uptake, including vaccine confidence, demographics and socioeconomic status.
- The survey is based on established behavioural theories, and is the Australian arm of the international iCARE survey which to date has collected global comparative information from over 90,000 respondents in 140 countries.
- Our survey was only available in English, which may have led to an underrepresentation of ethnic groups, and participation was voluntary, so our sample may be prone to selection bias from those with more interest or engagement in COVID-19.

## Introduction

The SARS-CoV-2 (COVID-19) pandemic has resulted in an estimated 211 million cases and 4.43 million deaths worldwide, including 44,028 cases and 981 deaths in Australia (1), as of August 2021. The R0 value has increased from 2-3 for the original Wuhan SARS-CoV-2 virus to 5-6 for the Delta variant of the SARS-CoV-2 virus currently dominating the world (2). Whilst vaccinated individuals can be infected with and transmit SARS-CoV-2, the vaccines reduce the likelihood for serious illness and subsequent hospitalisation and death by greater than 80% and 85% (3). Therefore, vaccinated populations are pivoting from the prevention of SARS-CoV-2 infections to instead accepting that the virus is endemic with the aim to minimise serious illness, hospitalisation, and death (4, 5)

Minimising serious illness, hospitalisations, and deaths will require high vaccination rates for SARS-CoV-2, and ongoing preventative health behaviours such as physical distancing and wearing face masks (6) to protect the unvaccinated (e.g. young children) and those in which the vaccine is less effective such as the immunocompromised.(7) It is now clear that combined behavioural strategies and vaccination (including boosters), are the pathway out of perpetual strict population level restrictions, which in Australia have included limiting gatherings, restricting education and work attendance, stay at home orders and closing both state and international borders (8, 9). Although these restrictions have been effective at reducing COVID-19 transmission and have prevented large numbers of deaths to date (10, 11), they come with serious economic, social and mental health costs that are unacceptable in the long term (8).

Australia is a country with a strong public health record, backed by high vaccination rates, high socioeconomic status, low population density and a universal free health care system. (12) These factors, alongside the strict policies including lockdowns, and Australia being an island nation, making it easier to secure borders, had contributed to Australia largely controlling the pandemic prior to the emergence of the Delta variant.(12) However, having a low SARS-CoV-2 vaccination rate, due to public concerns over the safety of the Astra Zeneca vaccine and a lack of supply of the mRNA vaccines, Australia has been susceptible to recent delta variant outbreaks.

Vaccine uptake is critical to the long-term management of the COVID-19 pandemic. To date, over 11% of the world’s population have received at least one dose of a SARS-CoV-2 vaccine (3). Vaccine supply and uptake needs to be accelerated globally to enhance protection against COVID-19.(13) Vaccine hesitancy and vaccine confidence are key determinants of vaccine uptake, and it is vital to understand factors associated with hesitancy. Vaccine confidence refers to the trust in the vaccines, the providers who administer it, and the science, processes, and policies behind it (14). Vaccine hesitancy is the sense of uncertainty in vaccines for a particular belief or reason.(14, 15) Vaccine hesitancy and reduced confidence may result in the refusal of, or delay in the acceptance of, a vaccination (16). Both vaccine hesitancy and confidence are complex and can be influenced by many determinants, (16) broadly grouped into three categories: 1) Contextual socio-politico-cultural factors, e.g., compatibility of vaccination with religious beliefs; 2) Individual and group influences, e.g., personal perception of the vaccine, or influences from the social and peer environment; and 3) Vaccine specific factors, e.g., issues directly related to the vaccine or vaccination such as the accelerated development of vaccines for SARS-CoV-2 may increase safety concerns in the population (8). Existing work on population intentions around the SARS-CoV-2 vaccines is emerging globally. A French study conducted early in the pandemic (March 2020) found that 26% of participants would not accept to receive a SARS-CoV-2 vaccine if it became available. (17) This was more prevalent amongst those in lower-income categories, young women and those older than 75 years of age. In the UK, 14% of participants in a study were unwilling to receive a vaccine, with 23% being unsure. (6) Similar to the French study, females and those from lower-income groups, reported being less likely to have a SARS-CoV-2 vaccine if available. (6)

The vaccine confidence index (VCI) consist of four questions to understand a person’s perceptions about if vaccines are safe, important, effective, and/or compatible with religious beliefs. (18). The VCI was developed following the identification of key drivers which influence the public’s confidence in vaccines. (14) Data has suggested approximately 1-in-5 Australians were hesitant regarding SARS-CoV-2 vaccines in the early stages of the COVID-19 pandemic in March/April 2020: with 14% to 24% respondents being unsure or unwilling to get a vaccine if available (19, 20).

This study identifies characteristics of Australians who intend or did not intend to get the vaccine in March 2021. Australia offers a unique case study to gain insights and inform mitigation strategies which could be applied globally. As attitudes towards the SARS-CoV-2 vaccine may vary over time, this new information will be able to inform current public health campaigns and policy (20, 21) and assist with effectively targeting those who currently have lower vaccine intentions. Hence here we aim to characterise the beliefs, intentions, and hesitancy of Australians towards vaccines generally (importance, safety and efficacy), and to SARS-CoV-2 specifically, to inform strategies to address this and increase uptake.

## Methods

This project is part of the Australian arm of the International COVID-19 Awareness and Responses Evaluation (iCARE) study, which is investigating people’s understanding, attitudes, beliefs, and actions towards the COVID-19 pandemic. (22) The Montreal Behavioural Medicine Centre, the lead institution, (23) has REB approval from the Comité d’éthique de la recherche du CIUSSS-NIM (Centre intégré universitaire de santé et de services sociaux du Nord-de-l’île-de-Montréal), approval#: 2020-2099/25-03-2020. This paper reports the analysis of the new vaccination questions asked in the third round of the Australian longitudinal survey (24); which comprised a national representative sample. The third round included new questions on attitudes towards the COVID-19 vaccination and intention to vaccinate against COVID-19 in Australia, therefore longitudinal comparison with earlier rounds (24) is not possible. Here we report the nationally representative cross-sectional analysis of respondents in this third survey conducted in early 2021. This project was approved by the Monash University Human Research Ethics Committee (#ID: 24449).

### Sampling

Survey respondents were recruited by an online sampling provider that sent out invitations between February 14^th^ and March 7^th^, 2021. By this time, Australia had recorded 28,947 COVID-19 cases with. variable virus impacts and policy approaches across states and a lack of national coordination. At a state level, Western Australia was lifting a lockdown (February 5^th^, 2021) and Victoria had entered a “circuit breaker,” 5-day lockdown having had more than 100 days in lockdown in 2020 (February 12th, 2012). The first public COVID-19 vaccinations were available on the 21^st^ of February, 2021. Electronic survey invitations were emailed to approximately 12,000 adults having a residential address in Australia, and briefly described the survey content, estimated survey duration, and a link to the online survey. The first page of the survey described the study, its purpose, and advised readers that continuing to the next page would be an indicator of consent to participate in the study. All participants who completed the online survey were reimbursed by ISO 26362 as per industry requirements. Representative sampling for key demographics of the Australian population was done using quota sampling for age, sex, and residential location (state/territory and remoteness area).

### Patient and public involvement

As part of the main iCARE study, there are several community collaborators who provide continual input into the development of the survey design, ensuring that the items are relevant and appropriately worded. For Australia, the survey was reviewed by the Monash Partners Consumer and Carer group and involved two members paid for their time to identify text that wasn’t clear or irrelevant to Australia, and recommend alternative wording and areas to clarify. Other community members and contacts of the researchers provided input into the timing to complete the survey, and subsequently this feedback resulted in the survey being shortened to reduce participant burden.

### Analysis Plan

Participant demographic data included residential postcode, which were mapped to the Australian Bureau of Statistics remoteness areas and socioeconomic index for areas (SEIFA). (25) Specifically, the index of relative socioeconomic disadvantage (IRSD) was applied and divided into five quintiles, from 1 (most disadvantaged) to 5 (most advantaged). Ethnicity information provided by participants was used to make two groupings of “Australian/New Zealand/UK” and everyone else. Descriptive statistics reported the participant demographics and attitudes for a series of vaccine-related questions including the VCI. (14, 18).

To characterise the beliefs, intentions and hesitancy of Australians towards vaccines generally and SARS-CoV -2 vaccines specifically, a series of univariate logistic regressions were done with dichotomous outcomes. Responses were dichotomised using the most extreme positive response e.g. “Always” vs other. To examine robustness, regressions were repeated by re-dichotomising outcomes to include the two most extreme responses instead of one. Unlike in similar analyses (18), our outcomes could not be examined using ordinal logistic regression because of low numbers in some response categories.

Possible predictors examined in the logistic regressions, included age, sex, essential worker status, belief that a participant is at high risk, residential area, flu vaccination status, education level, ethnicity, perceived income level, and IRSD quintile. Ethnicity data was missing for n=431 participants, therefore, these results were exploratory only. Responses to the VCI questions were also examined. All results are displayed as odds ratios, with 95% confidence intervals. Sensitivity analyses involved Bayesian logistic regression to enabled global comparisons with a recent Lancet publication. (18) and were conducted on the same outcome variables as in the logistic regressions. Normal priors (0,1) were set for each regression parameter and used 5,000 burn-in steps and 50,000 sampling iterations. Statistical analyses used STATA SE/v16. Significance level was set as <0.05.

## Results

There were 1166 survey respondents reported in this cross-sectional analysis. Response rate was approximately 10% for new participants and 60% for those in the longitudinal arm (24). Ages ranged from 18-90 years with a mean of 51.7 years (table 1), similar to the Australian population, apart from an overly represented group of participants aged 70 years or more. Sampling ensured a reasonable representativeness across sex, rurality and the three largest states (New South Wales, Victoria and Queensland). Education levels were similar to the Australian population. Less than half of participants (45%) were in full-or part-time work, lower than national statistics reported for the same time period (63%), and likely due to the overly represented 70+ age group. Fifteen percent reported being essential workers, with 7% healthcare workers. There was minimal missing data (table 1), except for ethnicity, with 50% respondents identifying as Australian/UK/NZ (n=580), 13% as other (n= 155) but 37% were missing (n=431).

**Table 1:**
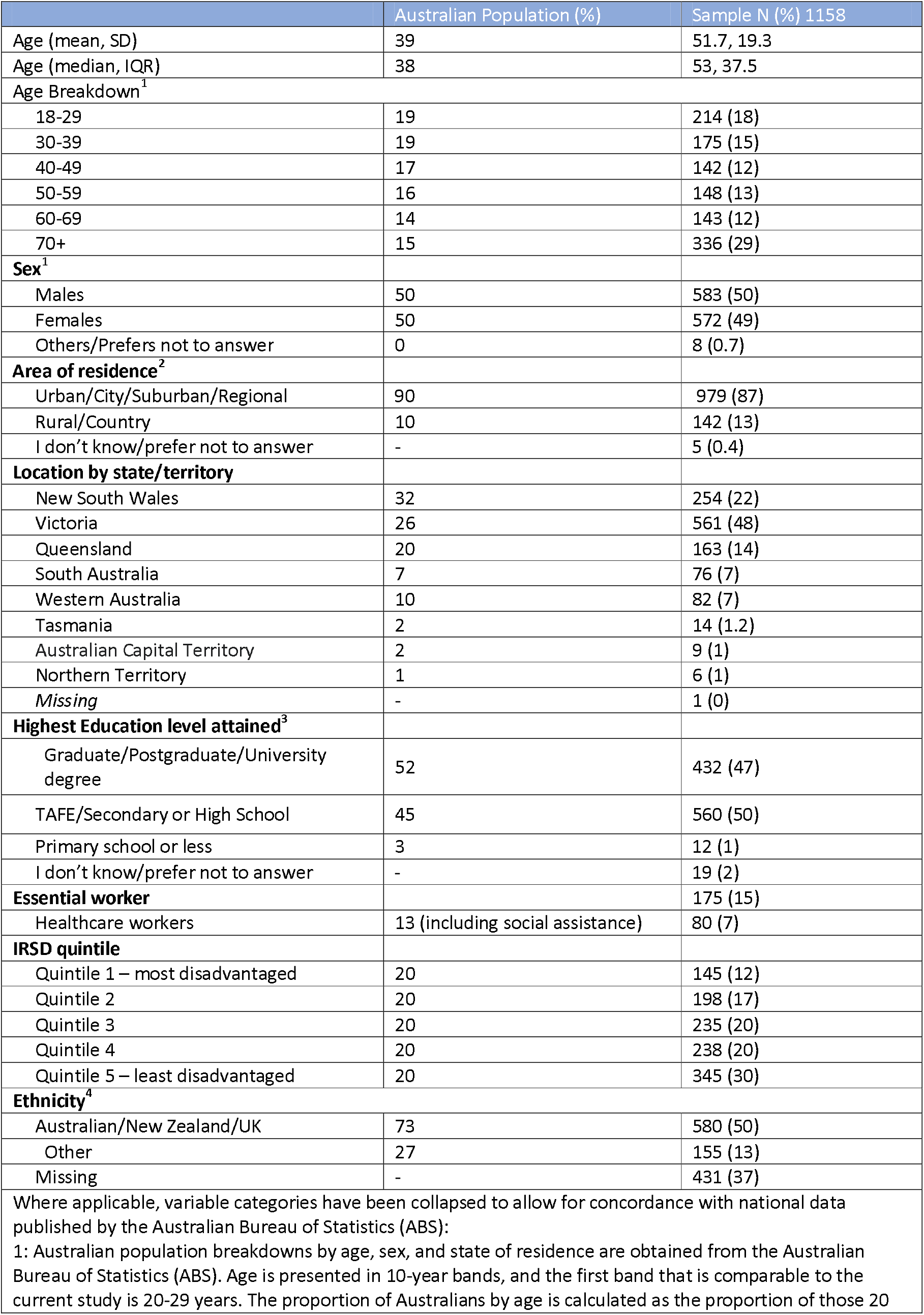
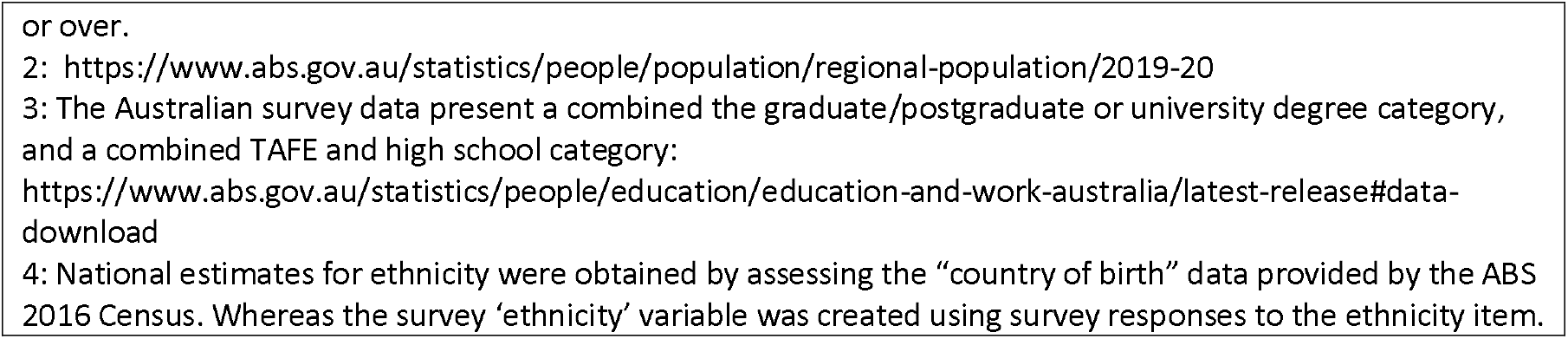
Participant demographics (n = 1166).

Sixty-five percent of participants generally accept routine vaccines for themselves or for their children, with 6% either rarely or never accepting vaccinations (table 2). At the time of this study, only 27 (2%) participants had already received at least one dose of a COVID-19 vaccination. The majority (78%) reported that they were likely to get the SARS-CoV-2 vaccine (table 2), and fifteen percent of all participants were either unlikely or very unlikely to get the SARS-CoV-2 vaccine. Seventy-two percent of our study cohort did not believe that there were at a high risk of being infected with COVID-19.

**Table 2:** Uptake intentions and attitudes on general vaccines and COVID-19 vaccine, and intent to continue engaging in virus-protecting behaviours (mask wearing, social distancing etc.) post COVID-19 vaccine.

|  | **N (%)** |
| --- | --- |
| **Had already received at least 1 dose of COVID-19 vaccine** | 27 (2) |
| **Likelihood of getting COVID-19 vaccine if it were available today** |  |
| Extremely likely | 597 (53) |
| Somewhat likely | 283 (25) |
| Unlikely | 88 (8) |
| Very unlikely | 83 (7) |
| I don't know/prefer not to answer | 80 (7) |
| **Generally accept vaccines for yourself or for your children** |  |
| Always | 736 (65) |
| Mostly | 232 (21) |
| Sometimes | 100 (9) |
| Rarely | 41 (4) |
| Never | 21 (2) |
| **Intent to continue engaging in virus-protecting behaviours (mask wearing, social distancing etc.) post vaccine** |  |
| Most of the time | 526 (47) |
| Some of the time | 343 (31) |
| Seldom | 94 (8) |
| Never | 54 (5) |
| I don't know/prefer not to answer | 95 (9) |
| **Seasonal flu vaccine over the last 5 years** |  |
| Every year | 511 (46) |
| 3-4 years | 163 (15) |
| 1-2 years | 202 (18) |
| Never | 218 (19) |
| I don't know/prefer not to answer | 27 (2) |

The VCI questions showed most Australians (>60%) strongly agreed on the safety, importance, and effectiveness of general vaccines (figure 1). Fifty-seven percent strongly agreed that general vaccines are compatible with their religious beliefs (figure 1). Approximately ten percent of participants did not know whether vaccines are safe or effective. (figure 1)

**Figure 1:**
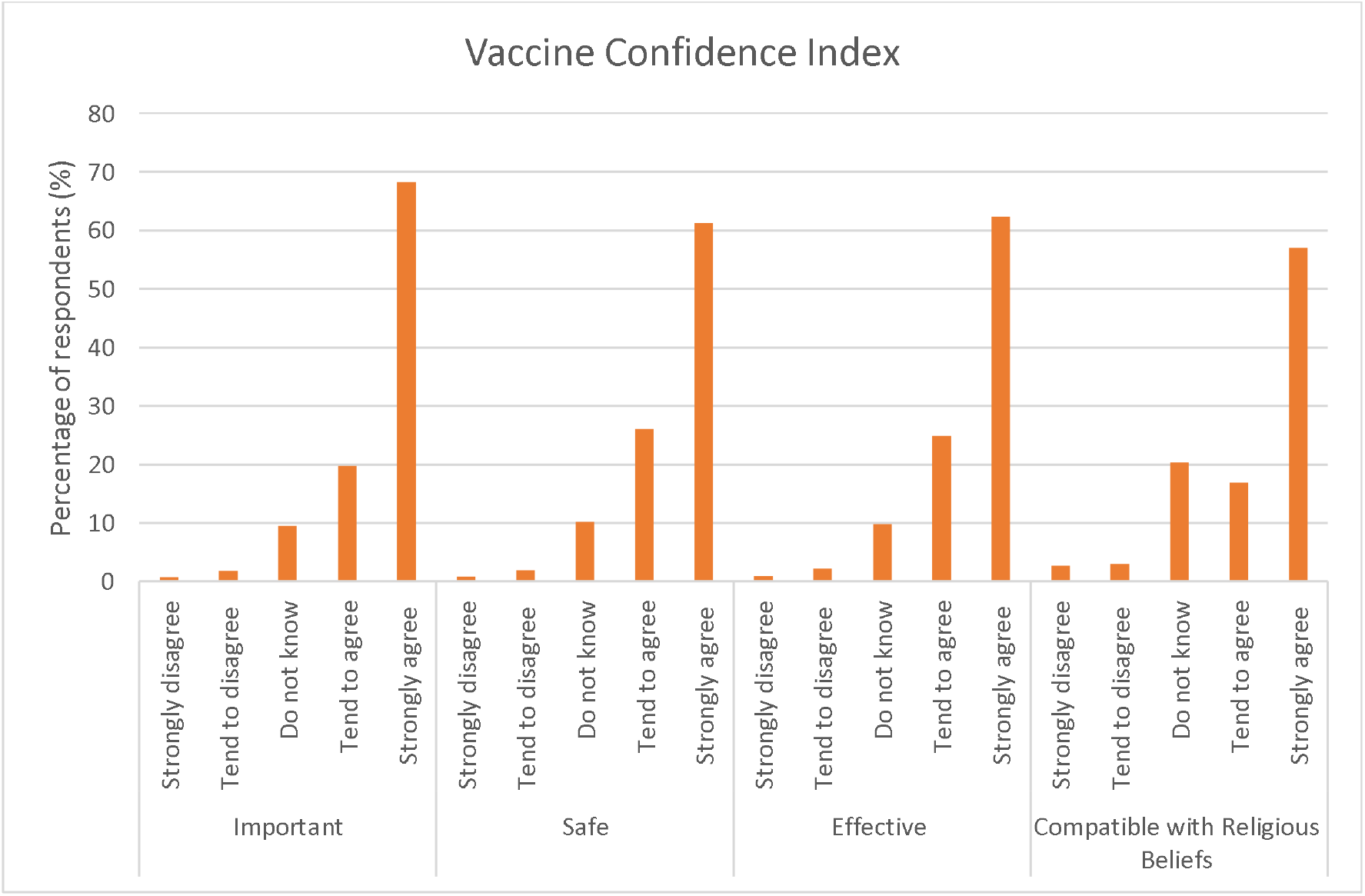
Vaccine Confidence Index: Responses to the questions about if general vaccines are safe, important, effective, and compatible with your religious beliefs.

### Predictors for vaccine uptake

Determinants that were similar for both general (Table 3) and SARS-CoV-2 vaccine uptake intention (Table 4) included:

**Table 4:** Factors reported by n=1,081 Australians that may influence intent to get the SARS-CoV-2 vaccine. *Combined ‘somewhat’ and ‘to a great extent’ responses. Influencing factors are ranked in descending order, from most likely to influence SARS-CoV-2 vaccine uptake to least likely.

| N (row %) | Combined strongest likelihood * | To a great extent | Somewhat | Very Little | Not at all | I don't know | Total |
| --- | --- | --- | --- | --- | --- | --- | --- |
| Having information that the vaccine is safe and unlikely to have any major long- term side effects | 921 (85) | 661 (61) | 260 (24) | 78 (7) | 50 (5) | 32 (3) | 1,081 |
| Having information that the vaccine is effective (i.e., provides a high degree of protection) | 913 (85) | 661 (61) | 252 (23) | 78 (7) | 58 (5) | 31 (3) | 1,080 |
| Knowing that getting vaccinated will help protect others around me | 858 (80) | 548 (51) | 310 (29) | 107 (10) | 72 (7) | 36 (3) | 1,073 |
| Trusting the company who developed the vaccine (Pfizer, Moderna, Sinopharm, etc.) | 839 (78) | 474 (44) | 365 (34) | 112 (10) | 75 (7) | 50 (5) | 1,076 |
| Receiving the vaccine dose(s) according to the manufacturers' instructions | 818 (76) | 505 (47) | 313 (29) | 122 (11) | 90 (8) | 42 (4) | 1,072 |
| Wanting to contribute to high population rates of vaccination to achieve 'herd immunity' | 791 (74) | 476 (44) | 315 (29) | 131 (12) | 101 (9) | 52 (5) | 1,075 |
| The convenience of getting the vaccine (e.g., requires little time, no need to travel far) | 772 (72) | 417 (39) | 355 (33) | 143 (13) | 118 (11) | 42 (4) | 1,075 |
| Getting a recommendation from my doctor to get vaccinated | 774 (72) | 438 (41) | 336 (31) | 163 (15) | 97 (9) | 37 (3) | 1,071 |
| Believing that I am high risk of getting COVID-19 or suffering severe complications | 729 (69) | 361 (34) | 368 (35) | 175 (17) | 119 (11) | 37 (3) | 1,060 |
| Learning that being vaccinated would allow me to attend public events (e.g., concerts, sporting events) or travel | 734 (68) | 422 (39) | 312 (29) | 179 (17) | 121 (11) | 39 (4) | 1,073 |
| Seeing more and more people getting the vaccine | 708 (66) | 335 (31) | 373 (35) | 191 (18) | 135 (13) | 34 (3) | 1,068 |
| Hearing that other people have positive attitudes towards the vaccine | 687 (64) | 306 (29) | 381 (36) | 195 (18) | 151 (14) | 35 (3) | 1,068 |
| Only needing one dose of the vaccine to be protected | 647 (61) | 302 (28) | 345 (32) | 203 (19) | 159 (15) | 56 (5) | 1,065 |
| Believing that getting vaccinated would reduce my worries and anxiety | 635 (60) | 273 (26) | 362 (34) | 225 (21) | 156 (15) | 43 (4) | 1,059 |
| Getting a recommendation from my employer to get vaccinated | 386 (52) | 158 (21) | 228 (31) | 163 (22) | 147 (20) | 44 (6) | 740 |

Higher likelihood of vaccine uptake was significantly associated with:

- Increasing age with Odds Ratio (OR) = 1.6 (95% Confidence Interval: 1.4-1.8) and 2.0 (95% CI: 1.8-2.3) for general and SARS-CoV-2 vaccine respectively; residing in the least disadvantaged areas SES quintile (OR = 2.1 (95% CI: 1.4-3.2) and 2.7 (95% CI: 1.5-3.4) for general and SARS-CoV-2 vaccines).
- Identifying as Australian/NZ/UK with an OR = 2.3 (95% CI: 1.6 - 3.3) and 1.9 (95% CI: 1.3 – 2.7) for general and SARS-CoV-2 vaccines; however, as noted there was much missing data for the ethnicity variable, therefore this result is considered exploratory only.
- Strong agreement with the VCI questions. For example, strong agreement with the statement “Vaccines are effective” had an OR = 14.6 (95% CI: 10.9 – 19.5) for general vaccine and 14.0 (95% CI: 10.4 – 18.9) for SARS-CoV-2 vaccine.

Lower likelihood of vaccine uptake was significantly associated with:

- Being a healthcare worker: With an OR of 0.5 (95%CI: 0.3 – 0.8) and 0.5 (95%CI: 0.3 – 0.8), for general and SARS-CoV-2 vaccines, respectively. However, this is exploratory only due to the small sample of healthcare workers and inability to delineate what worker type (e.g. allied health, medical, social worker, etc.)

There were no significant findings for educational level, perceived income or residential rurality. Differences between the general vaccines and the new COVID vaccines:

- There were no differences between the sexes for the likelihood of general vaccine uptake, whilst SARS-CoV-2 vaccine intention to uptake was significantly higher for men compared to women with OR of 1.37(95% CI: 1.08 – 1.72).

### Factors that might influence decisions to get the SARS-CoV-2 vaccine

Having information that the SARS-CoV-2 vaccine is safe (85%), effective (85%), will help protect people around the participant (80%), and trusting the company who developed the vaccine (78%) were reported to influence the participants somewhat or to a great extent to get vaccinated (Table 5). A doctor’s recommendation (72%) and convenience factors (72%) were also positive predictor variables for vaccine uptake. Other positive predictors include believing that the participant was at high risk of getting COVID-19 or suffering from severe complications (69%), increasing civil liberties (68%), and seeing others get vaccinated (66%).

### Sensitivity analyses

Bayesian regression analyses produced very similar results to initial logistic regression analyses. The regressions repeated with re-dichotomising outcomes to include the two most extreme responses instead of one, showed similar findings (supplementary table).

## Discussion

We examined the beliefs, intentions, and hesitancy of 1166 Australians towards vaccines in general and to the SARS-CoV-2 vaccine in a large, nationally representative cross-sectional analysis of a surveys in early 2021. Seventy-eight percent of all participants reported being likely to get the SARS-CoV-2 vaccine when it became available to them. Rates of both general vaccine uptake and SARS-CoV-2 vaccine uptake increased with age, believing that vaccines are safe and effective, and residing in the least disadvantaged socioeconomic region. Being male was associated with higher intentions to get the SARS-CoV-2 vaccine but had no statistically significant difference to general vaccine intention compared to females. There were no statistically significant differences in education level, perceived income level or rurality and rates of either general or SARS-CoV-2 vaccine acceptance. Strong influencing factors reported to convince people to uptake the SARS-CoV-2 vaccine were; knowing that the SARS-CoV-2 vaccine is safe and effective; trusting the vaccine producers; knowing it will help protect people close to them; recommendations from doctors to get vaccinated; and convenience getting the vaccine.

The following factors were identified as having more of an influence on vaccination rates, and hence could be inform public health policies and messaging to enhance vaccination rates. Having knowledge that the SARS-CoV-2 vaccine is safe and effective will encourage a large proportion of the study cohort to get vaccinated. These two factors are encompassed in the VCI and were recently examined in a large international study. (18) Together, they are likely to play the largest role in the uptake of the SARS-CoV-2 vaccine. Responsible, accurate reporting of the balance of risks and benefits in the media and social media is likely important to build trust in the vaccines and the companies that manufacture them.(26) Since trust in the vaccine companies is identified as a strong influencing factor in encouraging vaccination, this needs to be reaffirmed by focusing on the stringent regulatory processes the companies must adhere to, which can be conveyed in consistent and transparent public health messaging. Participants also indicated that knowing that the SARS-CoV-2 vaccine would protect those around them, was a significant factor influencing intention to vaccinate. Whilst those who are vaccinated can still transmit SARS-CoV2, transmission is decreased meaning family and friends are more protected, (27) which appeals to pro-social or altruistic attitudes, known to effectively increase vaccination rates.(28) Another key driver of vaccine uptake likelihood in our study was getting a recommendation from a doctor, aligned with previous immunisation programs, including in the H1N1 pandemic, and should be encouraged with the SARS-CoV-2 vaccine. (21) Medical professionals will benefit from consistent updated access to accurate information on the SARS-CoV-2 vaccine, countering non evidence based anti-vaccination messages, outlining benefits and risks, interpreting evidence as it emerges and personalising it to the individuals who seek care. (21, 29)

Convenience factors such as time needed or travel requirements to get vaccinated have also been identified as a strong influencing factor. This could translate to greater numbers of local vaccination sites in Australia, alongside the rollout of mass vaccination hubs, and of vaccinations to GP clinics, pharmacies, schools, and workplaces, already shown to increase the rate of other vaccinations including the annual influenza vaccine. (30) Here 68% of participants noted intention to get vaccinated if it offered them increased civil liberties, such as going to concerts or sporting events.

When choosing to get vaccinated, the perceived likelihood of infection, the prevalence and severity of the relevant disease are key in the decision making process. (31) In early 2021 in our study, 72% of all participants did not believe that they were at a high risk of getting COVID-19, likely reflecting the low numbers of infections, hospitalisations and deaths in Australia at that time. (32) Misinformation in the media also equated COVID-19 severity to that of the seasonal flu (33). These factors are likely to have presented obstacles to initial vaccination uptake in Australia, with participants perceived a higher risk of getting COVID-19 reporting 50% higher likelihood of getting vaccinated. Previous research on the SARS-CoV-2 vaccine, as well as vaccination research during the 2009 H1N1 pandemic echo our results. (29, 34). Leveraging anticipated regret, shown to be one of the strongest predictors for vaccine intention, could also be further explored to enhance SARS-CoV-2 vaccination rates. (29, 35) Consistent with other early surveys (36), we noted that men report the most willingness to receive a SARS-CoV-2 vaccine, however, this intention may not translate to gender differences in vaccination uptake (37).

Exploratory findings based on a small sample, suggested that healthcare workers and those not identifying themselves being from Australia/NZ/UK were less likely to accept both general and the SARS-CoV-2 vaccines. Considering the influence that healthcare workers have on the general population, their high exposure rates this presents a barrier to both effective vaccine uptake and to infection rates control. A 2021 review found an average of 23% (range: 4% to 72%) of healthcare workers reported vaccine hesitancy. The review also found that being male, older and a doctor were associated with higher rates of SARS-CoV-2 vaccine acceptance in healthcare workers. (38) The current study did not delineate between types of healthcare workers (e.g. doctors, nurses, allied health). Given pending policies around mandatory healthcare worker vaccination, the knowledge, attitudes and beliefs driving this behaviour needs further exploration. Additionally, our findings identified a higher rate of vaccine hesitancy in people who did not identify their ethnicity as Australian/New Zealanders or UK groups, consistent with past research in this and other vaccines. (29) However, the findings for both these high-risk groups need to be interpreted with caution due to the small sample size. Better data here could aid in further targeting policy-based communications and interventions.

Public health authorities need to provide transparent, easy to interpret information on the SARS-CoV-2 vaccines to the general population, as highlighted by Eastwood et al during the H1N1 pandemic. (39) This will aid in alleviating the confusion which may stem from misinformation present in the media and online networks. Furthermore, we echo the suggestions made in Seale et al., which includes tailoring messages and engaging community leaders in disseminating information about vaccines in culturally and linguistically diverse groups, with the known influence of social groups and community leaders of similar backgrounds. (40) For healthcare workers, engagement and education is important, given the important role they play in modelling health-promoting behaviour for the general public (37). Mandatory influenza vaccination is already in place for many healthcare workers in Australia, and mandatory SARS-CoV2 vaccination has been introduced for aged care workers and some jurisdictional healthcare workers with likelihood of scale up. (38). Finally, healthcare workers beliefs and attitudes to the SARS-CoV2 vaccine may reflect similar concerns to their broader community as seen in the UK with hesitancy being more frequent in non-white British healthcare workers, female sex, and younger age (41).

The strengths of our study include a large, generally representative sample across Australia and evidence based approaches including the vaccine confidence index. Limitations to our study include that this the survey was only available in English, which is likely to have reduced representation of ethnic groups. Internet access was required, which may account for the increased representation of those in the least disadvantaged quintile. Furthermore, since we rely on self-reported behaviour, there is the risk of a social desirability bias, with participants potentially over-reporting socially desirable traits in their responses and the voluntary nature of the survey makes it prone to a selection bias. (9, 42)

There is a paucity of studies on what influences people to consider taking the vaccine in Australia in 2021, where access to the SARS-CoV-2 vaccines is increasing, but still limited by age and occupation at the time of the survey. Since this survey, the rapid emergence of the highly transmissibility Delta variant, the major challenges of large scale, extended lockdowns escalating the imperative for rapid vaccination, and highlighting the importance of work in this field. Behavioural research such as the iCARE study can inform policymakers in understanding the public’s knowledge, attitudes, perceptions and beliefs towards the SARS-CoV-2 vaccine, which in turn drive their behaviours including vaccination and can aid with targeting public health messages. (21)

## Conclusion

Given the worldwide morbidity, hospitalisation, and death from COVID-19, the established safety and effectiveness of widely tested vaccines to prevent these complications, and the imperative to accelerate vaccination globally including in Australia, the results of this study on vaccine hesitancy are important. Here we show that vaccine safety, effectiveness, trust in the companies, and recommendations from doctors are important determinants of vaccine intentions. Further work to understand vaccine hesitancy in identified target groups including culturally and linguistically diverse groups and healthcare workers are important moving forward to support equity in vaccine uptake. This work can directly inform strategies to optimise communication and SARS-CoV-2 vaccine uptake, especially in Australia, now vital as the Delta variant takes a grip on the country.

## Supporting information

Strobe Checklist

## Data Availability

Data can be made available to approved researchers by contacting the corresponding author.

## Author Contributions

SB and KL led study conceptualisation. JE and JG contributed equally to this paper. JE and JG were responsible for the statistical analyses. JG and JE wrote the first draft of the paper. JB and HT are the senior authors and guarantors. All authors contributed to the development of the research question, study design in relation to the Australian data analysis, interpretation of the results, critical revision of the manuscript for important intellectual content, and approved the final version of the manuscript. JB attests that all listed authors meet authorship criteria and that no others meeting the criteria have been omitted.

## Licensing Statement

I, the Submitting Author has the right to grant and does grant on behalf of all authors of the Work (as defined in the below author licence), an exclusive licence and/or a non-exclusive licence for contributions from authors who are: i) UK Crown employees; ii) where BMJ has agreed a CC-BY licence shall apply, and/or iii) in accordance with the terms applicable for US Federal Government officers or employees acting as part of their official duties; on a worldwide, perpetual, irrevocable, royalty-free basis to BMJ Publishing Group Ltd (“BMJ”) its licensees and where the relevant Journal is co-owned by BMJ to the co-owners of the Journal, to publish the Work in BMJ Open and any other BMJ products and to exploit all rights, as set out in our <u>licence</u>.

---

**Table 2:**
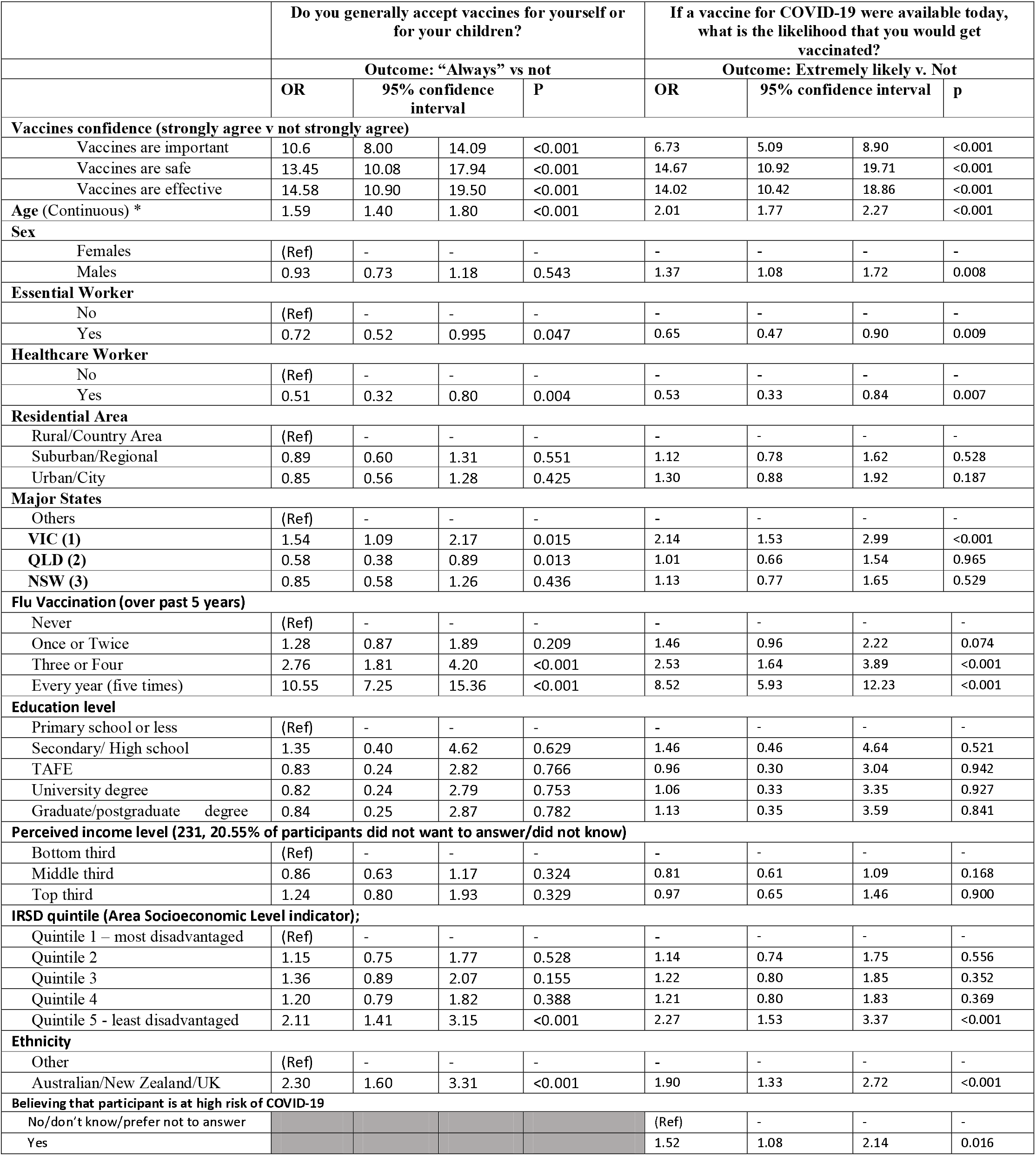
Vaccine uptake determinants: Univariate regression analyses with possible predictors that influence general vaccine uptake (left columns) and SARS-CoV-2 vaccine uptake(right columns). *Age variable is scaled to have a mean of 0 and unit standard deviation.**Ethnicity data was missing for n=431, therefore results for this variable are exploratory only.

