## Supplemental Table 1 for "Attitudes towards vaccines and intention to vaccinate against COVID-19: Implications for public health communications in Australia"

**Supplementary 1: Vaccine uptake sensitivity analysis**: Univariate regression analyses with possible predictors that influence general vaccine uptake (left columns) and SARS-CoV-2 vaccine uptake(right columns), using “Always/Mostly” and “Extremely/Somewhat likely” answer options. *Age variable is scaled to have a mean of 0 and unit standard deviation.**Ethnicity data was missing for n=431, therefore results for this variable are exploratory only.

|  | **Do you generally accept vaccines for yourself or for your children?** | | | | **If a vaccine for COVID-19 were available today, what is the likelihood that you would get vaccinated?** | | | | |
| --- | --- | --- | --- | --- | --- | --- | --- | --- | --- |
|  | **Outcome: “Always/Mostly” vs not** | | | | **Outcome: Extremely/somewhat likely v. not** | | | | |
|  | **OR** | **95% confidence interval** | | **P** | **OR** | **95% confidence interval** | | **p** | |
| **Vaccines confidence (strongly agree v not strongly agree)** | | | | | | | | | |
| Vaccines are important | 14.28 | 9.66 | 21.09 | <0.001 | 7.01 | 5.23 | 9.40 | <0.001 | |
| Vaccines are safe | 26.21 | 15.44 | 44.48 | <0.001 | 10.07 | 7.29 | 13.92 | <0.001 | |
| Vaccines are effective | 20.30 | 12.65 | 32.58 | <0.001 | 9.80 | 7.13 | 13.48 | <0.001 | |
| **Age** (Continuous) * | 1.53 | 1.31 | 1.80 | <0.001 | 1.60 | 1.39 | 1.84 | <0.001 | |
| **Sex** | | | | | | | | | |
| Females | (Ref) | - | - | - | - | - | - | - | |
| Males | 0.94 | 0.06 | 5.73 | 0.649 | 1.30 | 0.99 | 1.70 | 0.058 | |
| **Essential worker** | | | | | | | | | |
| No | (Ref) | - | - | - | - | - | - | - | |
| Yes | 0.91 | 0.19 | 0.93 | 0.034 | 0.86 | 0.59 | 1.25 | 0.426 | |
| **Healthcare worker** | | | | | | | | | |
| No | (Ref) | - | - | - | - | - | - | - | |
| Yes | 0.67 | 0.38 | 1.17 | 0.155 | 0.96 | 0.56 | 1.64 | 0.888 | |
| **Residential Area** | | | | | | | | | |
| Rural/Country Area | (Ref) | - | - | - | - | - | - | - | |
| Suburban/Regional | 0.95 | 0.56 | 1.62 | 0.844 | 1.01 | 0.66 | 1.55 | 0.956 | |
| Urban/City | 0.80 | 0.45 | 1.40 | 0.427 | 1.41 | 0.88 | 2.26 | 0.151 | |
| **Major States** |  |  |  |  |  |  |  |  | |
| Others | (Ref) | - | - | - | - | - | - | - | |
| **VIC (1)** | 1.45 | 0.93 | 2.27 | 0.100 | 1.40 | 0.95 | 2.05 | 0.089 | |
| **QLD (2)** | 0.68 | 0.41 | 1.14 | 0.140 | 0.69 | 0.44 | 1.10 | 0.117 | |
| **NSW (3)** | 0.90 | 0.56 | 1.46 | 0.672 | 0.94 | 0.62 | 1.45 | 0.796 | |
| **Flu Vaccination (over past 5 years)** | | | | | | | | | |
| Never | (Ref) | - | - | - | - | - | - | | - |
| Once or Twice | 2.06 | 1.32 | 3.21 | 0.001 | 2.38 | 1.58 | 3.59 | | <0.001 |
| Three or Four | 4.22 | 2.40 | 7.43 | <0.001 | 3.10 | 1.96 | 4.90 | | <0.001 |
| Every year (five times) | 18.62 | 10.22 | 33.93 | <0.001 | 10.26 | 6.73 | 15.63 | | <0.001 |
| **Education level** | | | | | | | | | |
| Primary school or less | (Ref) | - | - | - | - | - | - | | - |
| Secondary/ High school | 2.56 | 0.66 | 9.96 | 0.174 | 1.56 | 0.41 | 5.96 | | 0.518 |
| TAFE | 1.85 | 0.48 | 7.12 | 0.369 | 0.99 | 0.26 | 3.76 | | 0.989 |
| University degree | 1.46 | 0.38 | 5.57 | 0.579 | 1.04 | 0.27 | 3.93 | | 0.957 |
| Graduate/postgraduate degree | 2.58 | 0.66 | 10.12 | 0.174 | 1.31 | 0.34 | 5.01 | | 0.698 |
| **Perceived income level (231 (20.55%) of participants did not want to answer/did not know)** | | | | | | | | | |
| Bottom third | (Ref) | - | - | - | - | - | - | | - |
| Middle third | 1.30 | 0.86 | 1.96 | 0.210 | 1.40 | 0.98 | 2.00 | | 0.062 |
| Top third | 1.64 | 0.89 | 3.04 | 0.116 | 1.33 | 0.81 | 2.19 | | 0.256 |
| **IRSD quintile (Area Socioeconomic Level indicator)** | | | | | | | | | |
| Quintile 1 – most disadvantaged | (Ref) | - | - | - | - | - | - | | - |
| Quintile 2 | 0.81 | 0.46 | 1.42 | 0.458 | 1.25 | 0.78 | 1.99 | | 0.354 |
| Quintile 3 | 0.77 | 0.45 | 1.33 | 0.350 | 1.28 | 0.82 | 2.01 | | 0.281 |
| Quintile 4 | 0.83 | 0.48 | 1.43 | 0.496 | 1.39 | 0.88 | 2.19 | | 0.153 |
| Quintile 5 - least disadvantaged | 1.56 | 0.90 | 2.70 | 0.115 | 2.42 | 1.55 | 3.80 | | <0.001 |
| **Ethnicity** | | | | | | | | | |
| Other | (Ref) | - | - | - | - | - | - | | - |
| Australian/New Zealand/UK | 2.79 | 1.74 | 4.49 | <0.001 | 1.54 | 1.02 | 2.31 | | 0.04 |
| **Believing that participant is at high risk of COVID-19** | | | | | | | | | |
| No/don’t know/prefer not to answer |  |  |  |  | (Ref) | - | - | | - |
| Yes |  |  |  |  | 1.52 | 0.99 | 2.33 | | 0.054 |

**Supplementary 2: Standard vs. Bayesian logistic regression for SARS-CoV-2 vaccine uptake**: Univariate regression analyses with possible predictors that influence SARS-CoV-2 vaccine uptake, showing standard logistic regressions (left column) and Bayesian logistic regressions (right column). *Age variable is scaled to have a mean of 0 and unit standard deviation.

| **If a vaccine for COVID-19 were available today, what is the likelihood that you would get vaccinated?** | | | | | | | | |
| --- | --- | --- | --- | --- | --- | --- | --- | --- |
|  | Standard logit | | | | Bayesian logit | | | |
|  | OR | 95% confidence interval | | P | OR | 95% HPD interval | | p |
|  |  |  |  |  | Coef (median) |  |  |  |
| Vaccines are important (strongly agree v not strongly agree) | 6.73 | 5.09 | 8.90 | <0.001 | 6.45 | 4.86 | 8.35 | Significant- Does not cross 1 or 0 |
|  |  |  |  |  | 1.85 | 1.59 | 2.13 |  |
| Vaccines are safe (strongly agree v not strongly agree) | 14.67 | 10.92 | 19.71 | <0.001 | 13.76 | 10.07 | 18.05 | Significant |
|  |  |  |  |  | 2.61 | 2.32 | 2.90 |  |
| Vaccines are effective (strongly agree v not strongly agree) | 14.02 | 10.42 | 18.86 | <0.001 | 13.00 | 9.59 | 17.09 | Significant |
|  |  |  |  |  | 2.57 | 2.29 | 2.85 |  |
| Age (Continuous)* | 2.01 | 1.77 | 2.27 | <0.001 | 2.01 | 1.76 | 2.25 | Significant (does not cross 1 or 0) |
|  |  |  |  |  | 0.69 | 0.57 | 0.82 |  |
| Sex (Males v Females) | 1.37 | 1.08 | 1.72 | 0.008 | 1.38 | 1.06 | 1.70 | Significant (does not cross 1 or 0) |
|  |  |  |  |  | 0.32 | 0.08 | 0.55 |  |
| Essential Worker (Yes v No)  *Includes healthcare worker too | 0.65 | 0.47 | 0.90 | 0.009 | 0.67 | 0.47 | 0.89 | Significant |
|  |  |  |  |  | -1.32 | -2.13 | -0.58 |  |
| Healthcare worker (Yes v No) | 0.53 | 0.33 | 0.84 | 0.007 | 0.56 | 0.31 | 0.81 | Significant |
|  |  |  |  |  | -0.60 | -1.08 | -0.15 |  |
| Believe that participant is at high risk of COVID-19 (Yes v Not yes (No and I don’t know/prefer not to answer)) | 1.52 | 1.08 | 2.14 | 0.016 | 1.52 | 1.03 | 2.05 | Significant |
|  |  |  |  |  | 0.41 | 0.06 | 0.75 |  |
| COVID-19 app (extremely likely to get vs. not extremely likely to get) | 4.52 | 3.47 | 5.90 | <0.001 | 4.47 | 3.36 | 5.70 | Significant |
|  |  |  |  |  | 1.49 | 1.23 | 1.75 |  |
| Residential area | | | | | | | | |
| Rural/Country Area | (Ref) | - | - | - | - | - | - | - |
| Suburban/Regional | 1.12 | 0.78 | 1.62 | 0.528 | 1.13 | 0.75 | 1.55 | Not significant |
|  |  |  |  |  | 0.10 | -0.24 | 0.47 |  |
| Urban/City | 1.30 | 0.88 | 1.92 | 0.187 | 1.31 | 0.84 | 1.81 | Not significant |
|  |  |  |  |  | 0.25 | -0.13 | 0.63 |  |
| Flu Vaccination (over past 5 years) | | | | | | | | |
| Never | (Ref) | - | - | - | - | - | - | - |
| Once or Twice | 1.46 | 0.96 | 2.22 | 0.074 | 1.36 | 0.86 | 1.93 | Not significant |
|  |  |  |  |  | 0.29 | -0.12 | 0.68 |  |
| Three or Four | 2.53 | 1.64 | 3.89 | <0.001 | 2.34 | 1.41 | 3.30 | Significant |
|  |  |  |  |  | 0.83 | 0.42 | 1.24 |  |
| Every year (five times) | 8.52 | 5.93 | 12.23 | <0.001 | 7.82 | 5.25 | 10.43 | Significant |
|  |  |  |  |  | 2.04 | 1.71 | 2.38 |  |
| IRSD quintile (Socioeconomic status); Quintile 1 (Greater disadvantaged), Quintile 5 (Lack of disadvantage) | | | | | | | | |
| Quintile 1 (greater disadvantage) | (Ref) | - | - | - | - | - | - | - |
| Quintile 2 | 1.14 | 0.74 | 1.75 | 0.556 | 1.13 | 0.71 | 1.61 | Not significant |
|  |  |  |  |  | 0.11 | -0.31 | 0.50 |  |
| Quintile 3 | 1.22 | 0.80 | 1.85 | 0.352 | 1.21 | 0.79 | 1.70 | Not significant |
|  |  |  |  |  | 0.17 | -0.22 | 0.54 |  |
| Quintile 4 | 1.21 | 0.80 | 1.83 | 0.369 | 1.19 | 0.76 | 1.67 | Not significant |
|  |  |  |  |  | 0.16 | -0.24 | 0.53 |  |
| Quintile 5 (least disadvantage) | 2.27 | 1.53 | 3.37 | <0.001 | 2.23 | 1.47 | 3.08 | Significant |
|  |  |  |  |  | 0.78 | 0.41 | 1.15 |  |

**Supplementary 3: Standard vs. Bayesian logistic regression for general vaccine uptake**: Univariate regression analyses with possible predictors that influence general vaccine uptake, showing standard logistic regressions (left column) and Bayesian logistic regressions (right column). *Age variable is scaled to have a mean of 0 and unit standard deviation.

| **Do you generally accept vaccines for yourself or for your children?** | | | | | | | | |
| --- | --- | --- | --- | --- | --- | --- | --- | --- |
|  | Standard logit | | | | Bayesian logit | | | |
|  | OR | 95% confidence interval | | P | OR | 95% HPD interval | | p |
|  |  |  |  |  | Coef (median) |  |  |  |
| Vaccines are important (strongly agree v not strongly agree) | 10.6 | 8.00 | 14.09 | <0.001 | 10.17 | 7.49 | 13.07 | Significant |
|  |  |  |  |  | 2.31 | 2.04 | 2.59 |  |
| Vaccines are safe (strongly agree v not strongly agree) | 13.45 | 10.08 | 17.94 | <0.001 | 12.85 | 9.36 | 16.56 | Significant |
|  |  |  |  |  | 2.54 | 2.27 | 2.83 |  |
| Vaccines are effective (strongly agree v not strongly agree) | 14.58 | 10.90 | 19.50 | <0.001 | 13.91 | 9.93 | 17.92 | Significant |
|  |  |  |  |  | 2.62 | 2.33 | 2.91 |  |
| Age (Continuous)* | 1.59 | 1.40 | 1.80 | <0.001 | 1.59 | 1.40 | 1.79 | Significant |
|  |  |  |  |  | 0.46 | 0.34 | 0.59 |  |
| Sex (Males v Females) | 0.93 | 0.73 | 1.18 | 0.543 | 0.94 | 0.72 | 1.18 | Not significant (crosses 1 and 0) |
|  |  |  |  |  | -0.06 | -0.30 | 0.18 |  |
| Essential Worker (Yes v No)  *Includes healthcare worker too | 0.72 | 0.52 | 0.995 | 0.047 | 0.74 | 0.52 | 0.99 | Significant |
|  |  |  |  |  | -0.32 | -0.64 | -0.01 |  |
| Healthcare worker (Yes v No) | 0.51 | 0.32 | 0.80 | 0.004 | 0.54 | 0.32 | 0.80 | Significant |
|  |  |  |  |  | -0.64 | -1.08 | -1.18 |  |
| Believe that participant is at high risk of COVID-19 (Yes v Not yes (No and I don’t know/prefer not to answer)) | 1.40 | 0.97 | 2.01 | 0.070 | 1.41 | 0.92 | 1.93 | Not significant |
|  |  |  |  |  | 0.33 | -0.02 | -0.70 |  |
| COVID-19 app (extremely likely to get vs. not extremely likely to get) | 3.89 | 2.92 | 5.19 | <0.001 | 3.84 | 2.83 | 4.99 | Significant |
|  |  |  |  |  | 1.33 | 1.05 | 1.62 |  |
| Residential Area | | | | | | | | |
| Rural/Country Area | (Ref) | - | - | - | - | - | - | - |
| Suburban/Regional | 0.89 | 0.60 | 1.31 | 0.551 | 0.93 | 0.62 | 1.29 | Not significant |
|  |  |  |  |  | -0.08 | -0.46 | 0.28 |  |
| Urban/City | 0.85 | 0.56 | 1.28 | 0.425 | 0.89 | 0.56 | 1.25 | Not significant |
|  |  |  |  |  | -0.13 | -0.54 | 0.25 |  |
| Flu Vaccination (over past 5 years) | | | | | | | | |
| Never | (Ref) | - | - | - | - | - | - | - |
| Once or Twice | 1.28 | 0.87 | 1.89 | 0.209 | 1.23 | 0.80 | 1.69 | Not significant |
|  |  |  |  |  | 0.19 | -0.19 | 0.55 |  |
| Three or Four | 2.76 | 1.81 | 4.20 | <0.001 | 2.61 | 1.63 | 3.69 | Significant |
|  |  |  |  |  | 0.94 | 0.54 | 1.34 |  |
| Every year (five times) | 10.55 | 7.25 | 15.36 | <0.001 | 9.85 | 6.49 | 13.38 | Significant |
|  |  |  |  |  | 2.27 | 1.92 | 2.63 |  |
| IRSD quintile (Socioeconomic status); Quintile 1 (Greater disadvantaged), Quintile 5 (Lack of disadvantage) | | | | | | | | |
| Quintile 1 (greater disadvantage) | (Ref) | - | - | - | - | - | - | - |
| Quintile 2 | 1.15 | 0.75 | 1.77 | 0.528 | 1.15 | 0.71 | 1.66 | Not significant |
|  |  |  |  |  | 0.12 | -0.30 | 0.55 |  |
| Quintile 3 | 1.36 | 0.89 | 2.07 | 0.155 | 1.35 | 0.82 | 1.91 | Not significant |
|  |  |  |  |  | 0.28 | -0.12 | 0.70 |  |
| Quintile 4 | 1.20 | 0.79 | 1.82 | 0.388 | 1.20 | 0.77 | 1.71 | Not significant |
|  |  |  |  |  | 0.17 | -0.24 | 0.56 |  |
| Quintile 5 (least disadvantage) | 2.11 | 1.41 | 3.15 | <0.001 | 2.09 | 1.31 | 2.91 | Significant |
|  |  |  |  |  | 0.72 | 0.33 | 1.11 |  |
